# Australian children’s foot, ankle, and leg problems in primary care: a secondary analysis of the Bettering the Evaluation and Care of Health (BEACH) data

**DOI:** 10.1101/2022.02.12.22270840

**Authors:** Cylie M. Williams, Hylton B. Menz, Peter A. Lazzarini, Julie Gordon, Christopher Harrison

## Abstract

**Objectives:** To explore children’s foot, ankle and leg consultation patterns and management practices in Australian primary care.

**Design:** Cross-sectional, retrospective study

**Setting:** Australia Bettering the Evaluation and Care of Health program dataset.

**Participants:** Data were extracted for GPs and patients ≤18 years from April 2000 to March 2016 inclusive.

**Main outcome measures:** Demographic characteristics: sex, GP age groups (i.e. <45, 45-54, 55+ years), GP country of training, patient age grouping (0-4, 5-9, 10-14, 15-18 years), postcode, concession card status, Indigenous status, up to three patient encounter reasons, up to four encounter problems/diagnoses, and the clinical management actioned by the GP.

**Results:** Children’s foot, ankle or leg problems were managed at a rate of 2.05 (95% CI 1.99 to 2.11) per 100 encounters during 229,137 GP encounters with children. There was a significant increase in the rate of foot, ankle and leg problems managed per 100 children in the population, from 6.1 (95% CI: 5.3-6.8) in 2005-06 to 9.0 (95% CI: 7.9-10.1) in 2015-16. Management of children’s foot, ankle and leg problems were independently associated with male patients (30% more than female), older children (15-18 years were 7.1 times more than <1 years), male GPs (13% more) and younger GPs (<45 years of age 13% more than 55+). The top four most frequently managed problems were injuries (755.9 per 100,000 encounters), infections (458.2), dermatological conditions (299.4) and unspecified pain (176.3). The most frequently managed problems differed according to age grouping.

**Conclusions:** Children commonly present to GPs for foot, ankle, and leg problems. Presentation frequencies varied according to age. Unexpectedly, conditions presenting commonly in adults, but rarely in children, were also frequently recorded. This data highlights the importance of initiatives supporting contemporary primary care knowledge of diagnoses and management of paediatric lower limb problems to minimise childhood burden of disease.

**Article Summary:** *Strengths and limitations of this study:*

- This study examines the full spectrum of childhood foot, ankle, or leg presentations in primary care and how these are managed
- This study also provides information about how foot, ankle and leg GP presentations and management patterns differ as children get older
- This dataset provides a robust baseline on which future guidelines and implementation studies can measure the outcomes of practice change over time.
- This study may be limited by how GPs coded the presentation and management data

## Background

Childhood foot, ankle and leg concerns are thought to be common, but their prevalence and incidence vary widely according to age and are inconsistently reported. For example, the prevalence estimates for flexible flat foot vary from 2 to 44% of children ^1 2^, while the incidence of calcaneal apophysitis ranges from 0.37 to 0.60 per 100 person-years ^3^. These wide variations seem to depend on age, developmental stage, sporting participation or differences in epidemiological study setting. Similarly, little is known about the frequencies of conditions relating to the foot, ankle or leg in children that cause pain or functional impact or trouble their parents enough to result in families seeking management in primary care.

Key developmental stages in childhood present opportunities for optimal foot and leg condition management, particularly for conditions relating to musculoskeletal complaints, neurological conditions, or inflammatory disease. Early interventions for these higher burden conditions are important to initiate early to reduce long term disability and prevent chronic pain development. Conversely, delayed diagnosis, delayed access to care or provision of non-evidence informed care can be detrimental to long term outcomes ^4^, family burden ^5^ and permanent disability ^6^. Primary care providers are commonly the first contact for non-emergency health care. Therefore, understanding contemporary practice in primary care allows for improved focus for finite health care resources, training and guidelines, to improve health outcomes ^7^, reduce health care waste ^8^, and design effective public policies or prevention strategies to minimise long term impacts ^9^.

In Australia, primary care services are frequently provided by general practitioners (GPs) on a ‘fee for service’ model with fees primarily covered through Medicare, the Australian Government funded medical insurance scheme ^10^. Medicare also provides subsidies for other healthcare services including diagnostic imaging and pathology tests. The Pharmaceutical Benefits Scheme provides subsidies for prescribed medicines ^11^. GPs can also provide referrals to medical specialists for subsidised medical specialist care, such as to orthopaedic surgeons, and in limited circumstances (e.g. for chronic medical conditions) to subsidised allied health professional care, such as to podiatrists ^11^. Therefore, GP presentation and management data provides rich information about health problems in Australia.

Despite this, little is known about how GPs manage foot and leg problems in children in Australia, and even less about their management practices. It is important to know the frequencies of children’s foot, ankle, and leg problems and how commonly they present to GPs, as highly prevalent specific foot, ankle, or leg conditions in childhood may impact on health care costs now or in the future. Conservative estimates indicate that management of foot, ankle or leg conditions by GPs in Australia across all ages are estimated to be approximately A$255m per annum ^12^. Also unknown, is how many foot, ankle or leg conditions appear in childhood requiring medical care from GPs. To our knowledge, only four studies have examined presentations for primary care management in children that included lower limb presentations. These studies were in Spain, Australia, and the United Kingdom ^13-16^, yet only one of these studies provided data on all children between the ages of 0-18 years ^14^. Whilst studies have investigated the most frequent presenting conditions by children to GPs, they rarely delineate by body region such as foot, ankle and leg problems One Australian study reported data on all GP encounters by children aged between 0-17 years for any health condition and found frequent presentations for skin concerns and musculoskeletal concerns ^14^. Both skin and musculoskeletal concerns are two problems likely to include foot, ankle, or leg problems. However, there were no additional data on skin complaints relating to body region, and where musculoskeletal data according to body regions were explored in detail, lower limb concerns were managed at a rate between 0.62 to 5.33 per 100 children encounters. These insights warrant further detailed exploration given the frequency of presentations.

Therefore, the primary aim of this study was to determine the rate of GP encounters where foot, ankle, and leg (defined as below the knee) conditions were managed in children aged between 0-18 years. Secondary aims included exploring the patient and GP characteristics associated with these encounters, the rate of these encounters for children in different age groups, and the most frequent management practices for these encounters among the different age groups.

## Methods

### Dataset

Data were extracted from the Bettering the Evaluation and Care of Health (BEACH) study. This data set was constructed from a continuous, nationally representative study of GP clinical activity. Details of the BEACH study and methods of data coding and collection are published in detail elsewhere ^17^. Each year, a random sample of approximately 1,000 Australian GPs completed the BEACH study. These GPs recorded details from 100 consecutive patient encounters on structured paper data collection forms. Data captured included demographic characteristics such as patient’s age, sex, postcode, concession card status, Indigenous status, up to three patient reasons for the encounter, up to four problems/diagnoses managed during the encounter, and the clinical management actioned by the GP. Management strategies were coded, such as medications (supplied, advised, or prescribed), referrals for pathology or diagnostic imaging, referrals to other health professionals and any procedures provided by the GP during the clinical encounter. Pharmaceutical data were coded using the Coding Atlas of Pharmaceutical Substances (CAPS) ^18^ which maps to the Anatomical Therapeutic Chemical Classification System ^19^. All other data (including problems managed, non-pharmaceutical treatments, referrals and investigations) were coded using the Australian GP interface terminology known as ICPC-2 PLUS ^20^ by the BEACH research team, with automated classification to the International Classification of Primary Care, Version 2 (ICPC-2) ^21^.

Ethical approvals for ongoing BEACH dataset research purposes were provided by the Human Research Ethics Committee of the University of Sydney (Ref: 2012/130) and (from 2000 to 2010) the Ethics Committee of the Australian Institute of Health and Welfare. GPs provided implied informed consent to collect unidentified data about patients through return of information. Patients (or their parents or guardians) provided informed oral consent to the GP for their de-identified data to be included in the dataset.

### Participants and data elements

We initially identified all GP encounters for children and adolescents aged 0-18 years recorded from April 2000 until March 2016 within the dataset. We selected ICPC-2 PLUS terms that primarily related to problems specifically affecting the foot and ankle, but also included conditions that manifest below the knee (such as restless leg syndrome), dermatological conditions (such as tinea pedis), and congenital lower limb conditions (such as pes planus or genu valgum) through a previously reported expert consensus process (Supplementary dataset 1) ^12^.

### Statistical analysis

The BEACH dataset forms a single-stage cluster sample study design. The GP is the sampling unit, and the GP-patient encounter is the unit of inference. We used Survey procedures in SAS v9.4 to adjust for this cluster in all analyses. We initially extracted data from all encounters where the patient was aged 18 years or less. We then extrapolated the rate of management per encounter recorded in BEACH to the number of annual Medicare Benefits Scheduled GP items of services claimed for children to calculate the total number of foot/ankle/leg problems in children managed that year. We then divided this figure by the number of children in the population (Australian Bureau of Statistics population statistics) ^22^ to calculate the rate per child in the population. We calculated the rate of foot, ankle or leg problems managed per 100 encounters for children aged 0-18 years (with the age groups <1, 1-4, 5-9, 10-14 and 15-18 years) and analysed this by both GP and patient characteristics. Patient encounters were then grouped into comparable age clusters. Due to the low numbers of foot, ankle and leg problems managed at encounters, the <1 and 1-4 years ages were combined so that our final age groups were: 0-4, 5-9, 10-14, and 15-18 years. The most common types of foot, ankle and leg problems were examined and reported per 100,000 encounters for all ages, and for each age group. We also examined how these foot, ankle and leg problems were managed by GPs. Significant differences were determined through non-overlapping 95% confidence intervals (95% CI). This provided a conservative estimate of significance compared with the traditional alpha of <0.05 ^23^.

We used multivariate logistic regression to determine the GP and patient characteristics independently associated with a foot, ankle and leg problem being managed at an encounter. All GP and patient characteristics were included in the model.

## Results

### GP management rate for foot, ankle, and leg problems

Between April 2000 and March 2016, 15,472 GPs recorded 229,137 encounters meeting the extraction criteria (children aged 0-18 years), of which 4,694 were related to foot, ankle or leg problems. The foot, ankle, and leg problems were managed at a rate of 2.05 (95% CI 1.99 to 2.11) per 100 GP encounters with children. There was a significant increase in the rate of foot, ankle and leg problems managed per 100 children in the population, from 6.1 (95% CI: 5.3-6.8) in 2005-06 to 9.0 (95% CI: 7.9-10.1) in 2015-16 (Figure 1).

**Figure 1.**
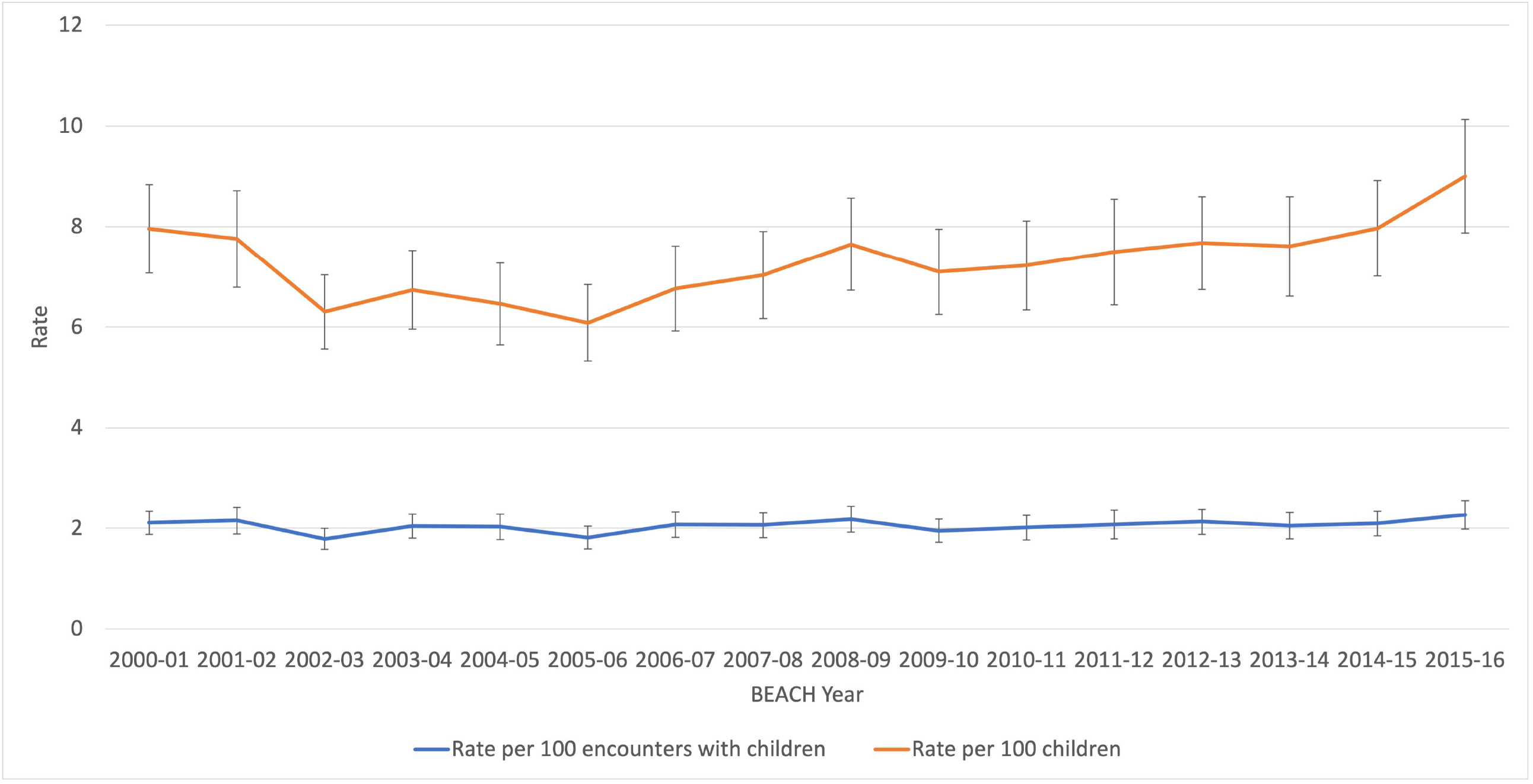
The management rate of children’s foot, ankle and leg problems managed by Australian GPs between April 2000 and March 2016 (aged 0-18 years). Blue line represents problems per 100 encounters, orange line represents problems per 100 children (Error bars = 95% CI).

### GP and child characteristics associated with management of foot, ankle, or leg problems

The highest rate of management was 4.64 (per 100 encounters) in the 10-14 years age group, the lowest was infants <1 year (0.44) (Table 1). After adjustment, male patients were 30% more likely to have afoot, ankle, or leg problem managed than their female peers at an encounter. Children in age groups 1-4, 5-9, 10-14 and 15-18 years were all more likely to receive care for foot ankle and leg conditions than children aged <1 year. Those aged 10-14 years were 10.2 times more likely than those aged <1 year. Those most disadvantaged were 8%more likely than those who were most advantaged. Male GPs were 13% more likely to provide care for a foot leg or ankle condition than female GPs. GPs aged <45 years were 13% more likely than those aged >55 years. Concession card status, being from a non-English speaking background, Indigenous status, practice location or GP country of training did not have a significant effect on whether a foot, ankle, and leg condition was managed.

**Table 1.**
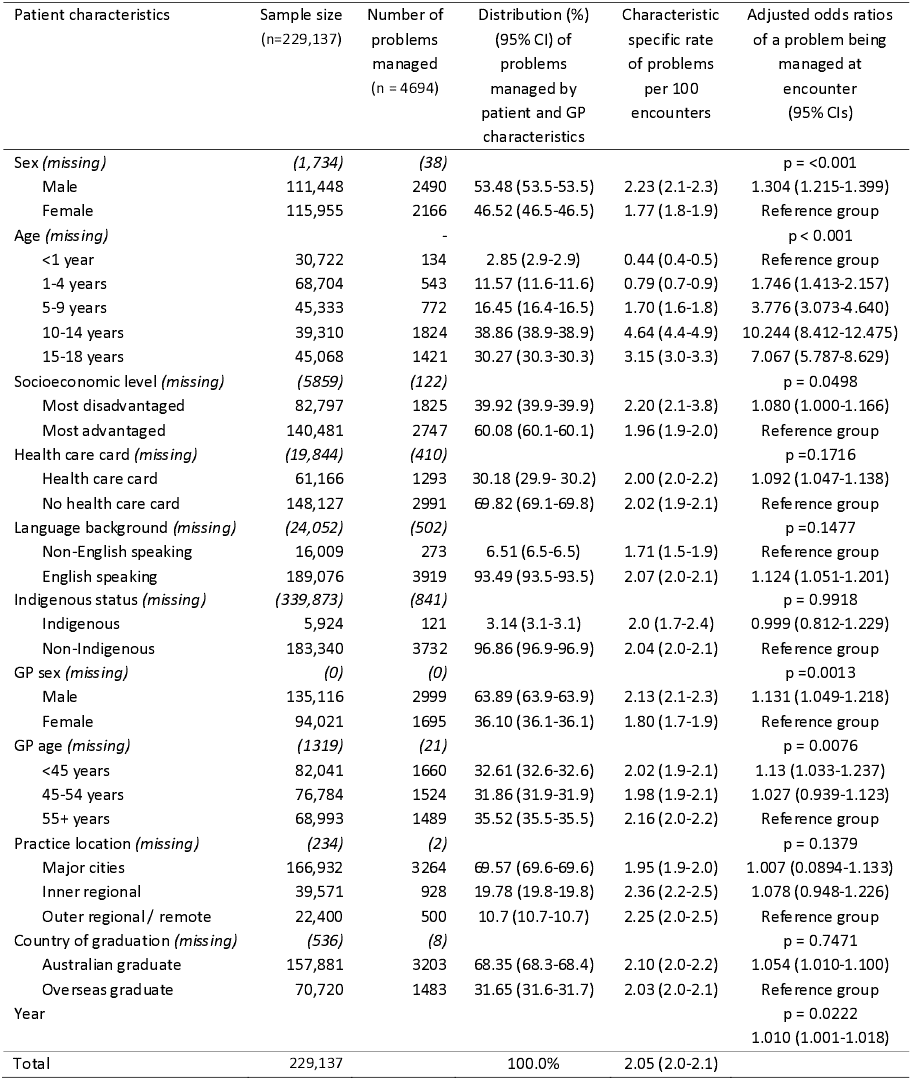
Child and GP specific management rate of foot/ankle/leg problems per 100 encounters, 2010-16.

| Patient characteristics | Sample size<br>(n=229,137) | Number of<br>problems<br>managed<br>(n = 4694) | Distribution (%)<br>(95% CI) of<br>problems<br>managed by<br>patient and GP<br>characteristics | Characteristic<br>specific rate<br>of problems<br>per 100<br>encounters | Adjusted odds ratios<br>of a problem being<br>managed at<br>encounter<br>(95% CIs) |
| --- | --- | --- | --- | --- | --- |
| Sex (*missing*) | (1,734) | (38) |  |  | p = <0.001 |
| Male | 111,448 | 2490 | 53.48 (53.5-53.5) | 2.23 (2.1-2.3) | 1.304 (1.215-1.399) |
| Female | 115,955 | 2166 | 46.52 (46.5-46.5) | 1.77 (1.8-1.9) | Reference group |
| Age (*missing*) | - | - |  |  | p < 0.001 |
| <1 year | 30,722 | 134 | 2.85 (2.9-2.9) | 0.44 (0.4-0.5) | Reference group |
| 1-4 years | 68,704 | 543 | 11.57 (11.6-11.6) | 0.79 (0.7-0.9) | 1.746 (1.413-2.157) |
| 5-9 years | 45,333 | 772 | 16.45 (16.4-16.5) | 1.70 (1.6-1.8) | 3.776 (3.073-4.640) |
| 10-14 years | 39,310 | 1824 | 38.86 (38.9-38.9) | 4.64 (4.4-4.9) | 10.244 (8.412-12.475) |
| 15-18 years | 45,068 | 1421 | 30.27 (30.3-30.3) | 3.15 (3.0-3.3) | 7.067 (5.787-8.629) |
| Socioeconomic level (*missing*) | (5859) | (122) |  |  | p = 0.0498 |
| Most disadvantaged | 82,797 | 1825 | 39.92 (39.9-39.9) | 2.20 (2.1-3.8) | 1.080 (1.000-1.166) |
| Most advantaged | 140,481 | 2747 | 60.08 (60.1-60.1) | 1.96 (1.9-2.0) | Reference group |
| Health care card (*missing*) | (19,844) | (410) |  |  | p = 0.1716 |
| Health care card | 61,166 | 1293 | 30.18 (29.9- 30.2) | 2.00 (2.0-2.2) | 1.092 (1.047-1.138) |
| No health care card | 148,127 | 2991 | 69.82 (69.1-69.8) | 2.02 (1.9-2.1) | Reference group |
| Language background (*missing*) | (24,052) | (502) |  |  | p = 0.1477 |
| Non-English speaking | 16,009 | 273 | 6.51 (6.5-6.5) | 1.71 (1.5-1.9) | Reference group |
| English speaking | 189,076 | 3919 | 93.49 (93.5-93.5) | 2.07 (2.0-2.1) | 1.124 (1.051-1.201) |
| Indigenous status (*missing*) | (339,873) | (841) |  |  | p = 0.9918 |
| Indigenous | 5,924 | 121 | 3.14 (3.1-3.1) | 2.0 (1.7-2.4) | 0.999 (0.812-1.229) |
| Non-Indigenous | 183,340 | 3732 | 96.86 (96.9-96.9) | 2.04 (2.0-2.1) | Reference group |
| GP sex (*missing*) | (0) | (0) |  |  | p = 0.0013 |
| Male | 135,116 | 2999 | 63.89 (63.9-63.9) | 2.13 (2.1-2.3) | 1.131 (1.049-1.218) |
| Female | 94,021 | 1695 | 36.10 (36.1-36.1) | 1.80 (1.7-1.9) | Reference group |
| GP age (*missing*) | (1319) | (21) |  |  | p = 0.0076 |
| <45 years | 82,041 | 1660 | 32.61 (32.6-32.6) | 2.02 (1.9-2.1) | 1.13 (1.033-1.237) |
| 45-54 years | 76,784 | 1524 | 31.86 (31.9-31.9) | 1.98 (1.9-2.1) | 1.027 (0.939-1.123) |
| 55+ years | 68,993 | 1489 | 35.52 (35.5-35.5) | 2.16 (2.0-2.2) | Reference group |
| Practice location (*missing*) | (234) | (2) |  |  | p = 0.1379 |
| Major cities | 166,932 | 3264 | 69.57 (69.6-69.6) | 1.95 (1.9-2.0) | 1.007 (0.0894-1.133) |
| Inner regional | 39,571 | 928 | 19.78 (19.8-19.8) | 2.36 (2.2-2.5) | 1.078 (0.948-1.226) |
| Outer regional / remote | 22,400 | 500 | 10.7 (10.7-10.7) | 2.25 (2.0-2.5) | Reference group |
| Country of graduation (*missing*) | (536) | (8) |  |  | p = 0.7471 |
| Australian graduate | 157,881 | 3203 | 68.35 (68.3-68.4) | 2.10 (2.0-2.2) | 1.054 (1.010-1.100) |
| Overseas graduate | 70,720 | 1483 | 31.65 (31.6-31.7) | 2.03 (2.0-2.1) | Reference group |
| Year |  |  |  |  | p = 0.0222 |
|  |  |  |  |  | 1.010 (1.001-1.018) |
| Total | 229,137 |  | 100.0% | 2.05 (2.0-2.1) |  |

### Rate of specific foot, ankle, and leg problems

Table 2 presents the child- and GP-specific management rate for the most common foot, ankle, and leg problem groups and specific conditions. The most frequently managed problem groupings were injuries (755.9 per 100,000 encounters), followed by infections (458.2) and dermatological conditions (299.4). The most frequent specific conditions were ankle sprains (310.3 per 100,000 encounters), ingrown toenails (272.3) or infected ingrown toenails (135.6), tinea or fungal skin infections (184.6), injuries to the foot/feet (76.4) and foot/feet pain (69.4). In general, management rates for problem groups and specific conditions increased with age until the 10 to 14 years age group, and then reduced in the 15-18 years group, except for the congenital problem groupings.

**Table 2.**
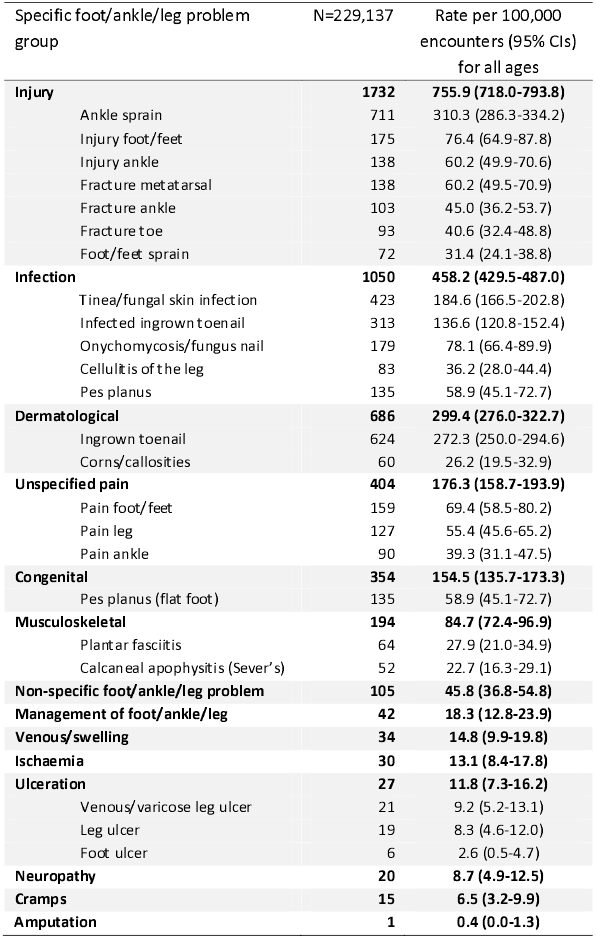
Management rate of foot/ankle/leg problem groups per 100,000 child encounters, 2000-16.

| Specific foot/ankle/leg problem group | N=229,137 | Rate per 100,000 encounters (95% CIs) for all ages |
| --- | --- | --- |
| **Injury** | **1732** | **755.9 (718.0-793.8)** |
| Ankle sprain | 711 | 310.3 (286.3-334.2) |
| Injury foot/feet | 175 | 76.4 (64.9-87.8) |
| Injury ankle | 138 | 60.2 (49.9-70.6) |
| Fracture metatarsal | 138 | 60.2 (49.5-70.9) |
| Fracture ankle | 103 | 45.0 (36.2-53.7) |
| Fracture toe | 93 | 40.6 (32.4-48.8) |
| Foot/feet sprain | 72 | 31.4 (24.1-38.8) |
| **Infection** | **1050** | **458.2 (429.5-487.0)** |
| Tinea/fungal skin infection | 423 | 184.6 (166.5-202.8) |
| Infected ingrown toenail | 313 | 136.6 (120.8-152.4) |
| Onychomycosis/fungus nail | 179 | 78.1 (66.4-89.9) |
| Cellulitis of the leg | 83 | 36.2 (28.0-44.4) |
| Pes planus | 135 | 58.9 (45.1-72.7) |
| **Dermatological** | **686** | **299.4 (276.0-322.7)** |
| Ingrown toenail | 624 | 272.3 (250.0-294.6) |
| Corns/callosities | 60 | 26.2 (19.5-32.9) |
| **Unspecified pain** | **404** | **176.3 (158.7-193.9)** |
| Pain foot/feet | 159 | 69.4 (58.5-80.2) |
| Pain leg | 127 | 55.4 (45.6-65.2) |
| Pain ankle | 90 | 39.3 (31.1-47.5) |
| **Congenital** | **354** | **154.5 (135.7-173.3)** |
| Pes planus (flat foot) | 135 | 58.9 (45.1-72.7) |
| **Musculoskeletal** | **194** | **84.7 (72.4-96.9)** |
| Plantar fasciitis | 64 | 27.9 (21.0-34.9) |
| Calcaneal apophysitis (Sever's) | 52 | 22.7 (16.3-29.1) |
| **Non-specific foot/ankle/leg problem** | **105** | **45.8 (36.8-54.8)** |
| **Management of foot/ankle/leg** | **42** | **18.3 (12.8-23.9)** |
| **Venous/swelling** | **34** | **14.8 (9.9-19.8)** |
| **Ischaemia** | **30** | **13.1 (8.4-17.8)** |
| **Ulceration** | **27** | **11.8 (7.3-16.2)** |
| Venous/varicose leg ulcer | 21 | 9.2 (5.2-13.1) |
| Leg ulcer | 19 | 8.3 (4.6-12.0) |
| Foot ulcer | 6 | 2.6 (0.5-4.7) |
| **Neuropathy** | **20** | **8.7 (4.9-12.5)** |
| **Cramps** | **15** | **6.5 (3.2-9.9)** |
| **Amputation** | **1** | **0.4 (0.0-1.3)** |

Table 3 outlines the management rate for foot, ankle, and leg problem groupings and specific conditions according to age group. The top three most frequently managed problem groupings were similar for all four age groups, with some exceptions in the younger age groups. Injuries (677.2 to 1835.7 per 100,000 encounters), infection (386.0 to 905.62), and dermatological conditions (101.5 to 877.6) were typically the top three in the older age groups (5 to 9 years, 10 to 14 years, and 15 to 19 years), although unspecified pain was the third most common problem group in those aged 5-9 years (257.1). For the youngest age group (0 to 4 years), the top three problem groupings were congenital (195.1), infection (191.1) and unspecified pain (57.3).

**Table 3.**
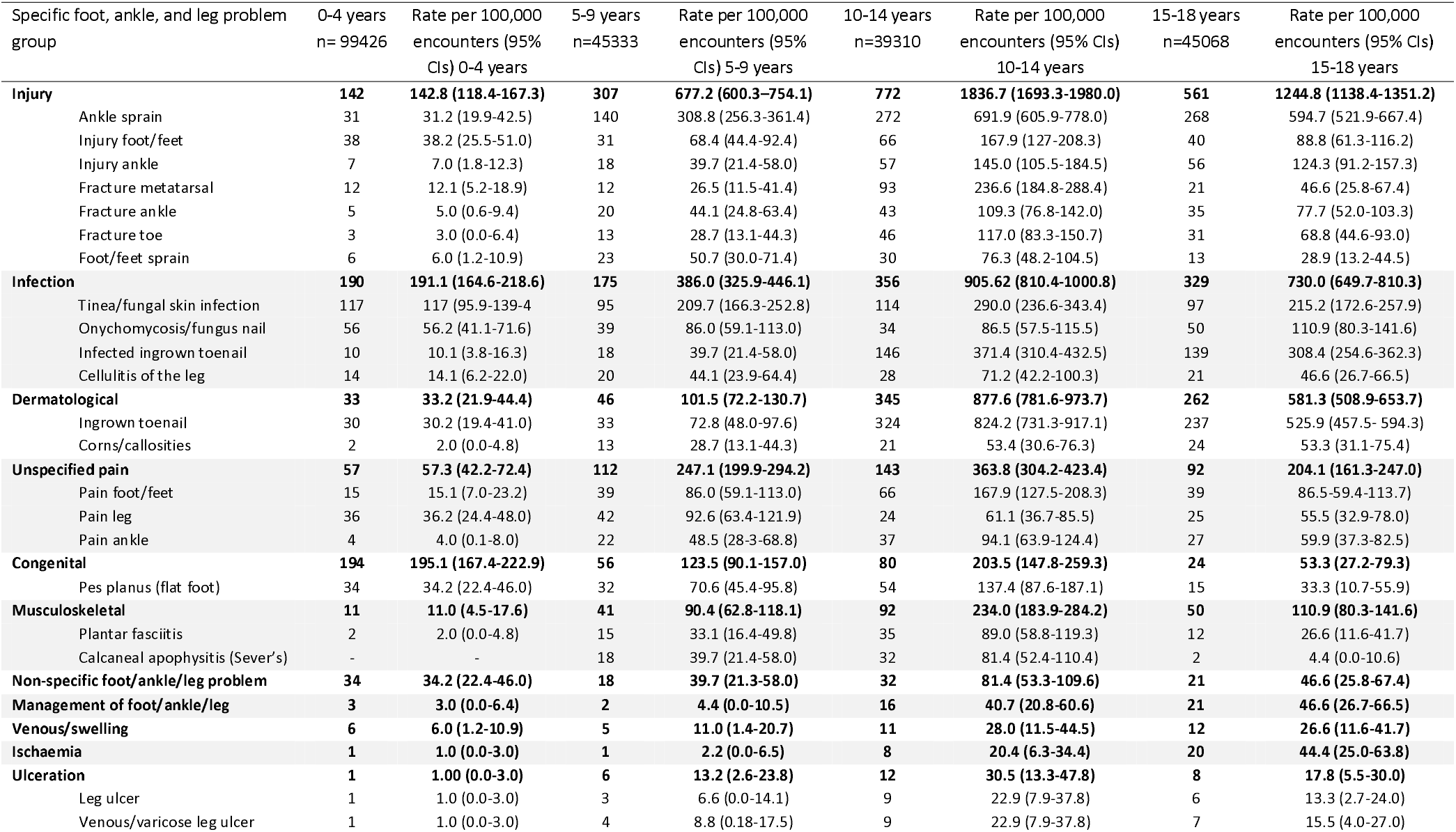
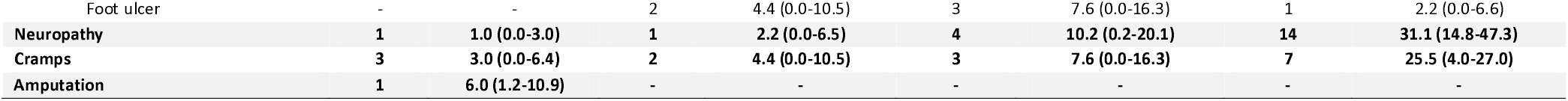
Management rate of paediatric foot/ankle/leg problem groups per 100,000 encounters, 2000-16 for age groupings

| Specific foot, ankle, and leg problem group | 0-4 years<br>n= 99426 | Rate per 100,000<br>encounters (95%<br>CIs) 0-4 years | 5-9 years<br>n=45333 | Rate per 100,000<br>encounters (95%<br>CIs) 5-9 years | 10-14 years<br>n=39310 | Rate per 100,000<br>encounters (95% CIs)<br>10-14 years | 15-18 years<br>n=45068 | Rate per 100,000<br>encounters (95% CIs)<br>15-18 years |
| --- | --- | --- | --- | --- | --- | --- | --- | --- |
| **Injury** | **142** | **142.8 (118.4-167.3)** | **307** | **677.2 (600.3-754.1)** | **772** | **1836.7 (1693.3-1980.0)** | **561** | **1244.8 (1138.4-1351.2)** |
| Ankle sprain | 31 | 31.2 (19.9-42.5) | 140 | 308.8 (256.3-361.4) | 272 | 691.9 (605.9-778.0) | 268 | 594.7 (521.9-667.4) |
| Injury foot/feet | 38 | 38.2 (25.5-51.0) | 31 | 68.4 (44.4-92.4) | 66 | 167.9 (127-208.3) | 40 | 88.8 (61.3-116.2) |
| Injury ankle | 7 | 7.0 (1.8-12.3) | 18 | 39.7 (21.4-58.0) | 57 | 145.0 (105.5-184.5) | 56 | 124.3 (91.2-157.3) |
| Fracture metatarsal | 12 | 12.1 (5.2-18.9) | 12 | 26.5 (11.5-41.4) | 93 | 236.6 (184.8-288.4) | 21 | 46.6 (25.8-67.4) |
| Fracture ankle | 5 | 5.0 (0.6-9.4) | 20 | 44.1 (24.8-63.4) | 43 | 109.3 (76.8-142.0) | 35 | 77.7 (52.0-103.3) |
| Fracture toe | 3 | 3.0 (0.0-6.4) | 13 | 28.7 (13.1-44.3) | 46 | 117.0 (83.3-150.7) | 31 | 68.8 (44.6-93.0) |
| Foot/feet sprain | 6 | 6.0 (1.2-10.9) | 23 | 50.7 (30.0-71.4) | 30 | 76.3 (48.2-104.5) | 13 | 28.9 (13.2-44.5) |
| **Infection** | **190** | **191.1 (164.6-218.6)** | **175** | **386.0 (325.9-446.1)** | **356** | **905.62 (810.4-1000.8)** | **329** | **730.0 (649.7-810.3)** |
| Tinea/fungal skin infection | 117 | 117 (95.9-139.4) | 95 | 209.7 (166.3-252.8) | 114 | 290.0 (236.6-343.4) | 97 | 215.2 (172.6-257.9) |
| Onychomycosis/fungus nail | 56 | 56.2 (41.1-71.6) | 39 | 86.0 (59.1-113.0) | 34 | 86.5 (57.5-115.5) | 50 | 110.9 (80.3-141.6) |
| Infected ingrown toenail | 10 | 10.1 (3.8-16.3) | 18 | 39.7 (21.4-58.0) | 146 | 371.4 (310.4-432.5) | 139 | 308.4 (254.6-362.3) |
| Cellulitis of the leg | 14 | 14.1 (6.2-22.0) | 20 | 44.1 (23.9-64.4) | 28 | 71.2 (42.2-100.3) | 21 | 46.6 (26.7-66.5) |
| **Dermatological** | **33** | **33.2 (21.9-44.4)** | **46** | **101.5 (72.2-130.7)** | **345** | **877.6 (781.6-973.7)** | **262** | **581.3 (508.9-653.7)** |
| Ingrown toenail | 30 | 30.2 (19.4-41.0) | 33 | 72.8 (48.0-97.6) | 324 | 824.2 (731.3-917.1) | 237 | 525.9 (457.5- 594.3) |
| Corns/callosities | 2 | 2.0 (0.0-4.8) | 13 | 28.7 (13.1-44.3) | 21 | 53.4 (30.6-76.3) | 24 | 53.3 (31.1-75.4) |
| **Unspecified pain** | **57** | **57.3 (42.2-72.4)** | **112** | **247.1 (199.9-294.2)** | **143** | **363.8 (304.2-423.4)** | **92** | **204.1 (161.3-247.0)** |
| Pain foot/feet | 15 | 15.1 (7.0-23.2) | 39 | 86.0 (59.1-113.0) | 66 | 167.9 (127.5-208.3) | 39 | 86.5-59.4-113.7) |
| Pain leg | 36 | 36.2 (24.4-48.0) | 42 | 92.6 (63.4-121.9) | 24 | 61.1 (36.7-85.5) | 25 | 55.5 (32.9-78.0) |
| Pain ankle | 4 | 4.0 (0.1-8.0) | 22 | 48.5 (28.3-68.8) | 37 | 94.1 (63.9-124.4) | 27 | 59.9 (37.3-82.5) |
| **Congenital** | **194** | **195.1 (167.4-222.9)** | **56** | **123.5 (90.1-157.0)** | **80** | **203.5 (147.8-259.3)** | **24** | **53.3 (27.2-79.3)** |
| Pes planus (flat foot) | 34 | 34.2 (22.4-46.0) | 32 | 70.6 (45.4-95.8) | 54 | 137.4 (87.6-187.1) | 15 | 33.3 (10.7-55.9) |
| **Musculoskeletal** | **11** | **11.0 (4.5-17.6)** | **41** | **90.4 (62.8-118.1)** | **92** | **234.0 (183.9-284.2)** | **50** | **110.9 (80.3-141.6)** |
| Plantar fasciitis | 2 | 2.0 (0.0-4.8) | 15 | 33.1 (16.4-49.8) | 35 | 89.0 (58.8-119.3) | 12 | 26.6 (11.6-41.7) |
| Calcaneal apophysitis (Sever's) | - | - | 18 | 39.7 (21.4-58.0) | 32 | 81.4 (52.4-110.4) | 2 | 4.4 (0.0-10.6) |
| **Non-specific foot/ankle/leg problem** | **34** | **34.2 (22.4-46.0)** | **18** | **39.7 (21.3-58.0)** | **32** | **81.4 (53.3-109.6)** | **21** | **46.6 (25.8-67.4)** |
| **Management of foot/ankle/leg** | **3** | **3.0 (0.0-6.4)** | **2** | **4.4 (0.0-10.5)** | **16** | **40.7 (20.8-60.6)** | **21** | **46.6 (26.7-66.5)** |
| **Venous/swelling** | **6** | **6.0 (1.2-10.9)** | **5** | **11.0 (1.4-20.7)** | **11** | **28.0 (11.5-44.5)** | **12** | **26.6 (11.6-41.7)** |
| **Ischaemia** | **1** | **1.0 (0.0-3.0)** | **1** | **2.2 (0.0-6.5)** | **8** | **20.4 (6.3-34.4)** | **20** | **44.4 (25.0-63.8)** |
| **Ulceration** | **1** | **1.00 (0.0-3.0)** | **6** | **13.2 (2.6-23.8)** | **12** | **30.5 (13.3-47.8)** | **8** | **17.8 (5.5-30.0)** |
| Leg ulcer | 1 | 1.0 (0.0-3.0) | 3 | 6.6 (0.0-14.1) | 9 | 22.9 (7.9-37.8) | 6 | 13.3 (2.7-24.0) |
| Venous/varicose leg ulcer | 1 | 1.0 (0.0-3.0) | 4 | 8.8 (0.18-17.5) | 9 | 22.9 (7.9-37.8) | 7 | 15.5 (4.0-27.0) |
| Foot ulcer | - | - | 2 | 4.4 (0.0-10.5) | 3 | 7.6 (0.0-16.3) | 1 | 2.2 (0.0-6.6) |
| **Neuropathy** | **1** | **1.0 (0.0-3.0)** | **1** | **2.2 (0.0-6.5)** | **4** | **10.2 (0.2-20.1)** | **14** | **31.1 (14.8-47.3)** |
| **Cramps** | **3** | **3.0 (0.0-6.4)** | **2** | **4.4 (0.0-10.5)** | **3** | **7.6 (0.0-16.3)** | **7** | **25.5 (4.0-27.0)** |
| **Amputation** | **1** | **6.0 (1.2-10.9)** | **-** | **-** | **-** | **-** | **-** | **-** |

The top three specific conditions were also similar for the older age groups (10-14 and 15-18 years) with ankle sprains (594.7 to 594.7), ingrown toenails (525.9 to 824.2) and infected ingrown toenails (308.4 to 371.4) being the top three in all those age groups. However, for the 0 to 4 years age group, the top three specific conditions were tinea or fungal skin infections (117.0), onychomycosis/fungal nail (56.2), and injuries to the foot/feet (39.2), while in the 5 to 9 years group, they were ankle sprains (308.8), tinea or fungal skin infections (209.7) and leg pain (92.6).

### Foot, ankle, and leg management strategies

Table 4 reports the most frequently used management strategies by GPs for foot, ankle, and leg problems. The top three most frequent action groupings were provision of medication (47.0 per 100 problems), counselling, advice, or education (25.4) and imaging (25.2). The most specific actions were referral for x-ray (22.7), prescription of antibiotics for systemic use (17.6), and prescription of analgesics (7.9).

**Table 4.** Management actions used by GPs for paediatric foot/ankle/leg problems, 2000-2016.

| Management action | n | Rate per 100 problems (95% CIs) |
| --- | --- | --- |
| **Medication (any)** | **2205** | **47.0 (45.2-48.8)** |
| *Antibiotics for systemic use* | 824 | 17.6 (16.4-18.7) |
| Cephalexin | 480 | 10.2 (9.2-11.1) |
| Flucloxacillin | 104 | 2.2 (1.8-2.7) |
| Dicloxacillin | 58 | 1.2 (0.9-1.6) |
| *Analgesics* | 370 | 7.9 (7.6-8.6) |
| Non-opioid analgesics | 311 | 6.6 (5.9-7.4) |
| Paracetamol | 277 | 5.9 (5.2-6.6) |
| Opioid analgesics | 59 | 1.3 (0.9-1.6) |
| Codeine/paracetamol | 55 | 1.2 (0.9-1.5) |
| Oxycodone | 4 | 0.1 (0.0-0.2) |
| Tramadol | 3 | 0.1 (0.0-0.1) |
| *Anti-inflammatory and antirheumatic products* | 228 | 4.9 (4.2-5.5) |
| Ibuprofen | 163 | 3.5 (2.9-4.0) |
| Meloxicam | 11 | 0.3 (0.1-0.4) |
| Diclofenac (oral) | 26 | 0.6 (0.3-0.8) |
| Diclofenac (topical) | 35 | 0.8 (0.5-1.0) |
| *Antifungals for dermatological use* | 354 | 7.5 (6.7-8.3) |
| Terbinafine (oral) | 27 | 0.6 (0.4-0.8) |
| Terbinafine (topical) | 67 | 1.4 (1.1-1.8) |
| Clotrimazole | 117 | 2.5 (2.0-2.9) |
| *Corticosteroids for dermatological use* | 120 | 2.6 (2.1-3.0) |
| Hydrocortisone | 16 | 0.3 (0.2-0.5) |
| **Procedures** | **997** | **21.2 (19.9-22.6)** |
| **Imaging** | **1185** | **25.2 (23.8-26.7)** |
| Ultrasound | 75 | 1.6 (1.2-2.0) |
| Xray | 1064 | 22.7 (21.3-24.0) |
| **Pathology** | **272** | **5.8 (4.6-7.0)** |
| Full blood count | 38 | 0.8 (0.5-1.1) |
| C-reactive protein | 13 | 0.3 (0.1-0.4) |
| Nail scraping/culture | 19 | 0.4 (0.2-0.6) |
| Skin swab/culture | 16 | 0.3 (0.2-0.5) |
| Fungal scraping/culture | 41 | 0.9 (0.6-1.1) |
| **Counselling/advice/education** | **1192** | **25.4 (24.0-26.8)** |
| **Referral** | **749** | **16.0 (14.8-17.1)** |
| Podiatrist | 182 | 3.9 (3.3-4.5) |
| Orthopaedic surgeon | 158 | 3.4 (2.8-3.9) |
| General surgeon | 65 | 1.4 (1.0-1.7) |
| Physiotherapist | 167 | 3.5 (3.0-4.1) |

Table 5 outlines the management strategies used according to age group. The top three most frequent management strategies were similar for the 5 to 9 years and 10 to 14 years age groups, although both the 0 to 4 years and 15 to 18 years exhibited different management patterns. For the 5 to 9 years and 10 to 14 years groups, the top three management strategies included medication prescription or advice (43.3 and 45.3 per 100 problems), imaging referral (27.2 and 30.7) and counselling, advice or education (25.8 and 27.7). In the 0 to 4 years group, the top three management strategies were medication prescription or advice (38.2), referral to another health professional (23.2) and counselling, advice, or education (21.2), whereas in the 15-18 years age group, it was medication prescription or advice (55.3), procedures, 24.4) and imaging referral (24.3). The top specific management strategies were similar for the 5 to 9 years and 10-14 years age groups. These were referrals for x-rays (24.6 to 28.4 per 100 problems), prescription of antibiotics for systemic use (11.1 to 20.4) and analgesics (7.7 to 9.5). The 0 to 4 age group top management strategies were referral for x-ray (15.2), antifungals for dermatological use (12.7) and prescriptions of antibiotics for systemic use (9.0), whereas, in the 15 to 18 years age group, the top three were prescription of antibiotics for systemic use (21.1), referral for x-ray (20.6) and analgesia (9.1).

**Table 5.** Management actions used by GPs for paediatric foot/ankle/leg problems, 2000-2016 for age groupings

| Management action | 0-4 years<br>n=677 | Rate per 100<br>problems (95% CIs)<br>for 0-4 years | 5-9<br>years<br>n=772 | Rate per 100 problems<br>(95% CIs) for 5-9 years | 10-14<br>years<br>n=1824 | Rate per 100<br>problems (95% CIs)<br>for 10-14 years | 15-18 years<br>n=1421 | Rate per 100<br>problems (95% CIs)<br>or 15-18 years |
| --- | --- | --- | --- | --- | --- | --- | --- | --- |
| **Medication (any)** | **259** | **38.2 (34.0-42.5)** | **334** | **43.3 (39.0-47.5)** | **826** | **45.3 (42.5-48.1)** | **786** | **55.3 (52.0-58.6)** |
| *Antibiotics for systemic use* | 61 | 9.0 (6.8-11.2) | 86 | 11.1 (8.9-13.4) | 373 | 20.4 (18.5-22.4) | 304 | 21.4 (19.2-23.6) |
| Cephalexin | 36 | 5.3 (3.6-7.0) | 45 | 5.8 (4.2-7.5) | 216 | 11.8 (10.3-13.3) | 183 | 12.9 (11.1-14.7) |
| Flucloxacillin | 6 | 0.9 (0.2-1.6) | 14 | 1.8 (0.9-2.8) | 48 | 2.6 (1.8-3.4) | 36 | 2.5 (1.7-3.4) |
| Dicloxacillin | - | - | - | - | 30 | 1.6 (1.0-2.2) | 28 | 2.0 (1.2-2.7) |
| *Analgesics* | 27 | 4.0 (2.5-5.5) | 73 | 9.5 (7.3-11.6) | 140 | 7.7 (6.4-9.0) | 130 | 9.1 (7.6-10.7) |
| Non-opioid analgesics | 26 | 3.8 (2.4-5.3) | 71 | 9.2 (7.0-11.4) | 123 | 6.7 (5.6-7.9) | 91 | 6.4 (5.1-7.7) |
| Paracetamol | 22 | 3.2 (1.9-4.6) | 62 | 8.0 (6.1-10.0) | 114 | 6.3 (5.1-7.4) | 79 | 5.6 (4.3-6.8) |
| Opioid analgesics | 1 | 0.1 (0.0-0.4) | 2 | 0.3 (0.0-0.6) | 17 | 0.9 (0.5-1.4) | 39 | 2.7 (1.9-3.6) |
| Codeine/paracetamol | 3 | 0.4 (0.00-0.9) | 1 | 0.1 (0.0-0.4) | 16 | 0.9 (0.4-1.3) | 35 | 2.5 (1.6-3.3) |
| Oxycodone | - | - | 1 | 0.1 (0.0-0.4) | 1 | 0.1 (0.0-0.2) | 2 | 0.1 (0.0-0.3) |
| Tramadol | - | - | - | - | 1 | 0.1 (0.0-0.2) | 2 | 0.1 (0.0-0.3) |
| *Anti-inflammatory and antirheumatic products* | 10 | 1.5 (0.6-2.4) | 32 | 4.1 (2.7-5.6) | 86 | 4.7 (3.7-5.7) | 100 | 7.0 (5.7-8.4) |
| Ibuprofen | 6 | 0.9 (0.2-1.6) | 31 | 4.0 (2.6-5.4) | 69 | 3.8 (2.9-4.7) | 57 | 4.0 (3.0-5.0) |
| Meloxicam | 1 | 0.1 (0.0-0.4) | - | - | 2 | - | 8 | 0.6 (0.2-1.0) |
| Diclofenac (oral) | 1 | 0.1 (0.0-0.4) | - | - | 10 | 0.5 (0.2-0.9) | 15 | 1.1 (0.5-1.6) |
| Diclofenac (topical) | 2 | 0.3 (0.0-0.7) | 5 | 0.6 (0.1-1.2) | 15 | 0.8 (0.4-1.2) | 13 | 0.9 (0.4-1.4) |
| *Antifungals for dermatological use* | 86 | 12.7 (10.2-15.3) | 73 | 9.5 (7.3-11.6) | 97 | 5.3 (4.2-6.4) | 98 | 6.9 (5.5-8.3) |
| Terbinafine (oral) | 5 | 0.7 (0.1-1.4) | 5 | 0.6 (0.1-1.2) | 10 | 0.5 (0.2-0.9) | 7 | 0.5 (0.1-0.9) |
| Terbinafine (topical) | 6 | 0.9 (0.2-1.6) | 19 | 2.5 (1.4-3.6) | 19 | 1.0 (0.6-1.5) | 23 | 1.6 (1.0-2.3) |
| Clotrimazole | 42 | 6.2 (4.4-8.0) | 21 | 2.7 (1.6-3.9) | 31 | 1.6 (1.1-2.3) | 8 | 1.6 (1.0-2.3) |
| *Corticosteroids for dermatological use* | 38 | 5.6 (3.8-7.4) | 30 | 3.8(2.5-5.3) | 26 | 1.4(0.9-2.0) | 26 | 1.8(1.1-2.6) |
| Hydrocortisone | 9 | 1.3 (0.5-2.2) | 4 | 0.5 (0.01-1.0) | 2 | - | 1 | 0.1 (0.0-0.2) |
| **Procedures** | **60** | **8.9 (1.2-6.4)** | **138** | **17.9 (14.8-20.9)** | **452** | **24.8 (22.5-27.1)** | **347** | **24.4 (21.8-27.0)** |
| **Imaging** | **107** | **15.8 (12.8-18.8)** | **236** | **30.7 (27.8-34.3)** | **496** | **27.2 (24.9-29.5)** | **346** | **24.3 (21.9-26.8)** |
| Ultrasound | 4 | 0.6 (0.01-1.2) | 10 | 1.3 (0.5-2.1) | 33 | 1.8 (1.2-2.4) | 25 | 2.0 (1.2-2.7) |
| Xray | 103 | 15.2 (12.3-18.2) | 219 | 28.4 (24.8-31.9) | 449 | 24.6 (1.1-26.8) | 293 | 20.6 (18.6-22.8) |
| **Pathology** | **41** | **6.1 (2.8-9.3)** | **61** | **7.9 (4.4-11.4)** | **86** | **4.7 (3.0-6.5)** | **84** | **5.9 (3.8-8.0)** |
| Full blood count | 6 | 0.8 (0.4-1.6) | 13 | 1.0 (0.8-2.6) | 10 | 0.5 (0.2-0.9) | 9 | 0.6 (0.2-1.0) |
| C-reactive protein | 4 | 0.6 (0.01-1.2) | 3 | 0.4 (0.0-0.8) | 3 | 0.2 (0.0-0.4) | 3 | 0.2 (0.0-0.4) |
| Nail scraping/culture | 2 | 0.3 (0.1-0.7) | 3 | 0.4 (0.0-0.8) | 4 | 0.2 (0.0-0.4) | 10 | 0.2 (0.3-0.4) |
| Skin swab/culture | 1 | 0.1 (0.0-0.4) | 1 | 0.1 (0.0-0.4) | 11 | 0.6 (0.2-1.0) | 3 | 0.1 (0-0.5) |
| Fungal scraping/culture | 8 | 1.2 (0.4-2.0) | 8 | 1.0 (0.3-1.8) | 16 | 0.9 (0.4-1.3) | 9 | 0.6 (0.2-1.0) |
| **Counselling/advice/education** | **179** | **21.2 (19.9-22.6)** | **214** | **27.7 (24.4-31.1)** | **471** | **25.8 (23.6-28.0)** | **328** | **23.1 (20.7-25.4)** |
| **Referral** | **157** | **23.2 (19.8-26.6)** | **112** | **14.5 (11.9-17.2)** | **277** | **15.2 (13.4-16.9)** | **203** | **14.3 (12.4-16.2)** |
| Podiatrist | 18 | 2.7 (1.3-4.1) | 39 | 5.1 (3.5-6.6) | 85 | 4.7 (3.7-3.1) | 40 | 2.8 (2.0-3.7) |
| Orthopaedic surgeon | 51 | 7.5 (5.5-9.5) | 23 | 3.0 (1.8-4.2) | 44 | 2.4 (1.7-3.1) | 40 | 2.8 (1.9-3.7) |
| General surgeon | 1 | 0.1 (0.0-0.4) | 3 | 0.2 (0.0-0.8) | 32 | 1.8 (1.1-2.4) | 29 | 2.0 (1.3-2.8) |
| Physiotherapist | 26 | 3.8 (2.4-5.3) | 21 | 2.7 (1.5-3.9) | 61 | 3.3 (2.5-4.2) | 59 | 4.2 (3.1-5.2) |

## Discussion

This study was one of the first to investigate the national management of children’s foot, ankle, and leg conditions by GPs. Findings suggest Australian GPs commonly manage children’s lower limb problems, and more frequently in males and older children. Injury, infection, and dermatological conditions presented most frequently to GPs across all ages and medications were the most frequently used management strategy. The frequency of specific problems managed, and the management strategies used, varied across the different age groupings, such as differing rates of congenital problems, or differing prescription or advice of medications. GPs also commonly provided counselling, advice and education for all ages, an appropriate management strategy for concerned parents, and a common first stage management strategy for many benign congenital, or undefined foot, ankle, or leg problems or while undergoing further testing to refine diagnosis ^24^.

Children from more disadvantaged socioeconomic areas had a significantly higher GP management rate of foot, leg and ankle conditions than their peers in more advantaged areas. This presentation is consistent with other studies on children’s healthcare in countries with socialised medicine, and reflects a complex interaction between health literacy of parents or the knowledge or financial ability for parents to seek health care information or alternate care providers without a GP recommendation, such as seeing a podiatrist or physiotherapist for their children’s foot, ankle or leg concerns ^25 26^.

Foot, ankle, and leg problems differed across age groupings and in general, increased with age. The presentations patterns extracted from this dataset related to foot, ankle, or leg concerns potentially reflect the different key skeletal and developmental stages. Younger children presented more with congenital lower limb concerns, while older children presented with more dermatological (e.g., tinea or ingrown toenails) or injury (e.g., ankle sprain) concerns. These presentation patterns align with key gross motor or developmental stages and may also align with the different health professional referral patterns. For example, there were higher numbers of congenital foot problems in younger children, and more frequent referrals to orthopaedic surgeons than in older age groups. In contrast, injuries were more common in older children, who were referred more often to podiatrists and physiotherapists. These patterns may reflect the more emergent nature of ensuring right timed surgical care at key osseus stages versus providing rehabilitation during injury recovery or individualised skin or nail care advice.

Injury was the primary problem managed in all ages. This may be due to different mechanisms of injuries occurring across childhood such as those occurring in the playground, or during social or organised sport ^27-30^. Despite how injuries may have occurred, common management strategies extracted from this dataset included frequent medical imaging. Ultrasound and x-rays were the most common imaging methods, with fewer ordered than frequency of injury presentations. This suggests conservative and judicious imaging referrals, and potential use of injury imaging referral guidelines, such as the Ottawa Ankle Rules ^31^.

Antibiotic stewardship and pain management medication strategies elicited from this dataset also mirror prescribing guidelines established for general practice relating to childhood presentations involving the lower limb for the timeframe data were extracted ^32^. For example, at the time of data collection, cephalexin was commonly prescribed in a suspension for children to treat mild skin infections (e.g., cellulitis) while narrow spectrum antibiotics such as flucloxacillin and dicloxacillin were the recommended antibiotics for infected skin relating to infected ingrown toenail presentations ^32^. Similarly, the use of non-opioid pain medications exceeded opioid prescriptions, consistent with recommended actions for pain management practices ^32^. We did not undertake direct comparisons between the problem managed and corresponding management strategy during this analysis; however, these patterns suggest that medication management practices align with best practice clinical guidelines.

Contrary to this, it was surprising to see fewer musculoskeletal conditions recorded within the dataset, despite epidemiological studies finding that 12% of children report or seek care for leg or foot pain relating to specific musculoskeletal conditions ^33^. The low frequency rates we observed within this dataset may be related to several factors. The most likely reason is how these problems were recorded by the GP. Underpinning how problems were recorded may be limited knowledge about less common foot, ankle or leg conditions, lower presentation rates of foot, ankle or leg conditions to GPs compared with hospital outpatients, the single point data collection used in the BEACH dataset that captures a problem as a symptom with as yet unknown diagnosis (e.g., waiting test results for confirmation) and relevant management guidelines of the time.

The low frequency of musculoskeletal concerns recorded within this dataset may also reflect different health literacy in parents and its association with not seeking care from GPs, or misdiagnosis. One potential example of this was the frequency of plantar fasciitis diagnoses recorded across younger ages (33.1 per 100,000 in the 5-9 year group and 89.0 per 100,000 in the 10 to 14 year group). Plantar fasciitis is rarely reported in contemporary paediatric orthopaedic literature, and if diagnosed on ultrasound, associated with being an older and highly athletic adolescent than the ages in this dataset ^34^. Heel pain in older children is more likely to be calcaneal apophysitis. This diagnosis was recorded as 39.1 per 100,000 encounters in 5-9 year grouping and 81.4 in 10-14 year grouping, and at a less frequent rate than plantar fasciitis in the 10-14 year age grouping, despite this being the age when foot apophyseal injuries are most prevalent ^3^. Other conditions also resulting in childhood plantar heel pain include inflammatory disease, infection (including osteomyelitis) or post-viral joint pain, all presenting more commonly than plantar fasciitis in younger age groupings ^35^. Management strategies of these heel pain conditions differ significantly, making it imperative for timely and accurate diagnosis to minimise health care wastage, and prevent development of chronic pain ^24^.

Recently there has been a global call to action on improving primary care diagnosis and assessment of musculoskeletal conditions in childhood to minimise misdiagnosis and reduce the development of disability and chronic pain ^36^. Simple assessments and screening tools have been implemented to support general practice, such as the paediatric Gait, Legs and Spine (p-GALS) screen ^37^, and free online generic health professionals training to improve paediatric musculoskeletal condition diagnoses ^38^. These resources have been developed in acknowledgement of limited exposure to paediatric musculoskeletal conditions during medical training ^39^, less common presentations in childhood compared to other childhood complaints such as ear infections or upper respiratory tract infection leading to low confidence in diagnostic skills of musculoskeletal conditions ^40^, knowledge deficits of the types of common paediatric musculoskeletal presentations ^41^, and serious long term consequences of some musculoskeletal conditions missed or misdiagnosed ^36^. Our findings of GPs reporting unspecified pain or conditions known to be more prevalent in adults than children suggests that Australian GPs may require additional support to diagnose and manage musculoskeletal conditions in childhood. Future research may include development of guidelines and supporting models of care for children’s foot, ankle, or leg problems to determine if these improve health outcomes, reduce the progressive nature of many musculoskeletal conditions and pain syndromes and if these are cost-effective.

This study is the first to our knowledge, to examine the full spectrum of childhood foot, ankle, or leg presentations in primary care and how these are managed. The data extracted from a large and representative sample of Australian GPs provides an extensive snapshot of practice to guide future directions for education, guideline development and models of care for childhood foot, ankle, or leg conditions. A limitation of this study is the historical nature of the data and that education, practice and models of care may have evolved between the 2016 end date of BEACH and data analysis. Known paediatric model of care and referral changes in some Australian state and territories occurred in late 2015 ^42^, which may have resulted in improved management of conditions through several guidelines, recommended assessments and when to refer to orthopaedic surgeons for several specific musculoskeletal conditions. Regardless, this dataset of encounters and management strategies provides a robust baseline on which future guidelines and implementation studies can measure the outcomes of practice change over time.

## Conclusion

Childhood foot, ankle and leg conditions are a common reason parents bring their children to a GP in Australia. Frequencies of presentations vary according to developmental stage with potential under reporting of musculoskeletal conditions. Future studies should consider how to support GPs in managing childhood musculoskeletal conditions to minimise disability and development of chronic pain. These actions have the potential to reduce long term burden of disease.

## Data Availability

The data that support the findings of this study are available from BEACH, but restrictions apply to the availability of these data, which were used under license for the current study, and so are not publicly available. Data are however available from the authors upon reasonable request and with permission of BEACH.

## Declarations

### Ethics approval and consent to participate

Not applicable.

### Consent for publication

Not applicable.

### Author Contributions

CW and PL conceived this study with authors HBM, JG and CH contributing to data extraction plan. CH undertook data extraction and analysis; CW developed the first draft of the manuscript with all authors providing critical review. All authors approved the final draft for submission

## Acknowledgements

We would like to thank the GPs who participated in the BEACH project for their generosity. We would also like to thank the BEACH team for collecting the data over the years. The BEACH study was conducted by the University of Sydney, and from April 1998 to March 2000, this was in collaboration with the Australian Institute of Health and Welfare.

